# Beyond Nominal Awareness: Mapping the Literacy to Action Chasm in Adolescent HPV Vaccination Across Sub-Himalayan India

**DOI:** 10.64898/2026.09.06.26362000

**Authors:** Debankur Chakraborty, S. Vanshika

## Abstract

**Background:** Cervical cancer is a major cause of death among Indian women, even though the HPV vaccine and routine screening can prevent it. Himachal Pradesh launched a state-funded HPV vaccination drive in early 2026. This study looks at what adolescent schoolgirls know about cervical cancer, whether they understand the vaccine, and what influences their choices.

**Methods:** We surveyed 516 female students across seven schools (five government and two private) in Bilaspur and Shimla districts of Himachal Pradesh between May and July 2026 (93.8% response rate). The survey covered knowledge of cervical cancer, HPV, vaccine benefits, vaccination status, fears about infertility, and willingness to get vaccinated if advised by a doctor.

**Results:** While 71.9% (371/516) of girls had heard of cervical cancer, only 47.3% (244/516) knew it begins in the cervix or lower uterus, 76.0% (392/516) knew no symptoms, and only 27.5% (142/516) recognized that it can be prevented (*p* < 0.001). Similarly, 61.6% (318/516) had heard of the HPV vaccine, but only 41.9% (216/516) had heard of HPV itself (*p* < 0.001), and only 50.0% (258/516) knew the vaccine prevents infection. Only 12.8% (66/516) had received at least one dose, and just two girls (0.4%) had finished all required doses. When asked if they would get vaccinated on a doctor’s advice, 45.3% (234/516) wanted more information first, 34.1% (176/516) refused, and 20.5% (106/516) agreed. Three out of four girls (75.6%, 390/516) were concerned or uncertain about whether the vaccine causes infertility. Only 3.5% (18/516) had heard of a Pap smear.

**Conclusion:** The findings demonstrate a pronounced decoupling between nominal awareness and actionable prevention literacy among adolescent schoolgirls in Himachal Pradesh. The predominant driver of vaccine hesitation was an unmet informational requirement rather than active ideological refusal. School-based immunization campaigns must incorporate structured, age-appropriate educational curriculum addressing the need of vaccination and infertility misconceptions prior to injection sessions.

## Introduction

Cervical cancer is a major global health challenge, causing around 660,000 cases and 350,000 deaths every year, with over 85% occurring in low- and middle-income countries [1], [2]. In India, it is the second most common cancer among women, leading to roughly 127,000 new cases and nearly 80,000 deaths annually [1], [3]. Long-term infection with high-risk human papillomavirus (HPV) especially types 16 and 18 causes more than 95% of all cervical cancers [2], [4]. HPV vaccines protect against these infections, and routine screenings detect early cell changes before they turn into cancer.

The World Health Organization (WHO) aims for 90% of girls to be vaccinated by age 15, 70% of women to be screened by ages 35 and 45, and 90% of women with cervical disease to be treated by 2030 [1], [4]. Reaching this goal requires vaccinating girls in early adolescence before they are exposed to the virus. In India, vaccination numbers have remained low due to procurement logistics, funding challenges, and limited public health communication [5], [6]. A 2024 review of 27 Indian studies found low overall HPV knowledge and large regional differences in uptake [5], [6]. Structured educational sessions in schools help young people understand and accept the vaccine [7]. At this age, adolescent choices are shaped by personal knowledge, conversations with peers and parents, and community beliefs, especially concerns about future childbearing [8], [9].

Most surveys only ask whether people have “heard of” a disease or vaccine. This does not show whether they understand what it does. A student might recognize the term “HPV vaccine” while mistakenly believing it cures colds, treats advanced cancer, or causes infertility. This study looks beyond surface-level awareness to examine what students know about cervical cancer, HPV transmission, vaccine purpose, actual uptake, infertility rumours, and screening awareness.

## Methods

### Study Design and Setting

This cross-sectional survey was conducted between May and July 2026 across seven schools in Bilaspur and Shimla districts, Himachal Pradesh. Bilaspur covers semi-urban and rural foothill valleys, while Shimla includes both the urban state capital and mountain settlements. Five government schools and two private schools participated.

### Participants

Schools were selected based on administrative permission and operational access. The study was conducted alongside Project Satya, an educational non-profit organization. Out of 550 eligible female students, 20 were absent and 14 left their forms incomplete, leaving 516 validated surveys for analysis (93.8% response rate).

### Survey Questions

Trained interviewers administered structured questionnaires using simple, everyday language across five core topics:

1. **Demographics:** Age group (<15, 15–17, 18–21, and ≥22 years) and schooling level.
2. **Cervical Cancer Knowledge:** General awareness, tumour location, symptoms, and perceived preventability.
3. **HPV Knowledge:** Awareness of the virus, its role as the main cause of cervical cancer, and transmission routes.
4. **Vaccine Awareness and Uptake:** Hearing about the vaccine, understanding its preventive purpose, and personal vaccination status.
5. **Decisions and Misconceptions:** Response to a doctor’s recommendation, beliefs about infertility, and Pap smear awareness.

### Statistical Analysis

All analyses used the 516 completed responses as the denominator. Proportions are reported with 95% Wilson confidence intervals (CIs). Gaps between related areas (such as awareness vs. preventability) were evaluated using absolute risk differences (ARD) and two-sample proportion z-tests (*p* < 0.05).

## Results

### Demographic Breakdown

Most participants (91.7%) were minors under 18 years of age, and girls under 15 formed the largest group (57.2%). Nearly two-thirds (64.0%) were enrolled in middle or high school classes.

**Table 1.** Participant Characteristics (N = 516)

| *Characteristic* | *Count*<br>*(n)* | *Percentage*<br>*(%)* | *95%*<br>*Wilson*<br>*CI* |
| --- | --- | --- | --- |
| **Age** |  |  |  |
| *Under 15 years* | 295 | 57.2% | 52.9%–61.4% |
| *15–17 years* | 178 | 34.5% | 30.5%–38.7% |
| *18–21 years* | 36 | 7.0% | 5.1%–9.5% |
| *22 years and older* | 7 | 1.4% | 0.7%–2.8% |
| **School Level** |  |  |  |
| *Middle / High School* | 330 | 64.0% | 59.7%–68.0% |
| *Senior Secondary /*<br>*Pre-University* | 186 | 36.0% | 32.0%–40.3% |

### What Students Know About Cervical Cancer

Although 71.9% of girls had heard of cervical cancer, only 47.3% knew it begins in the cervix or lower uterus. Three out of four (76.0%) could not name a single symptom, and only 27.5% knew the disease is preventable—a 44.4% drop compared to general awareness (z= 14.26, *p* < 0.001).

**Table 2.**
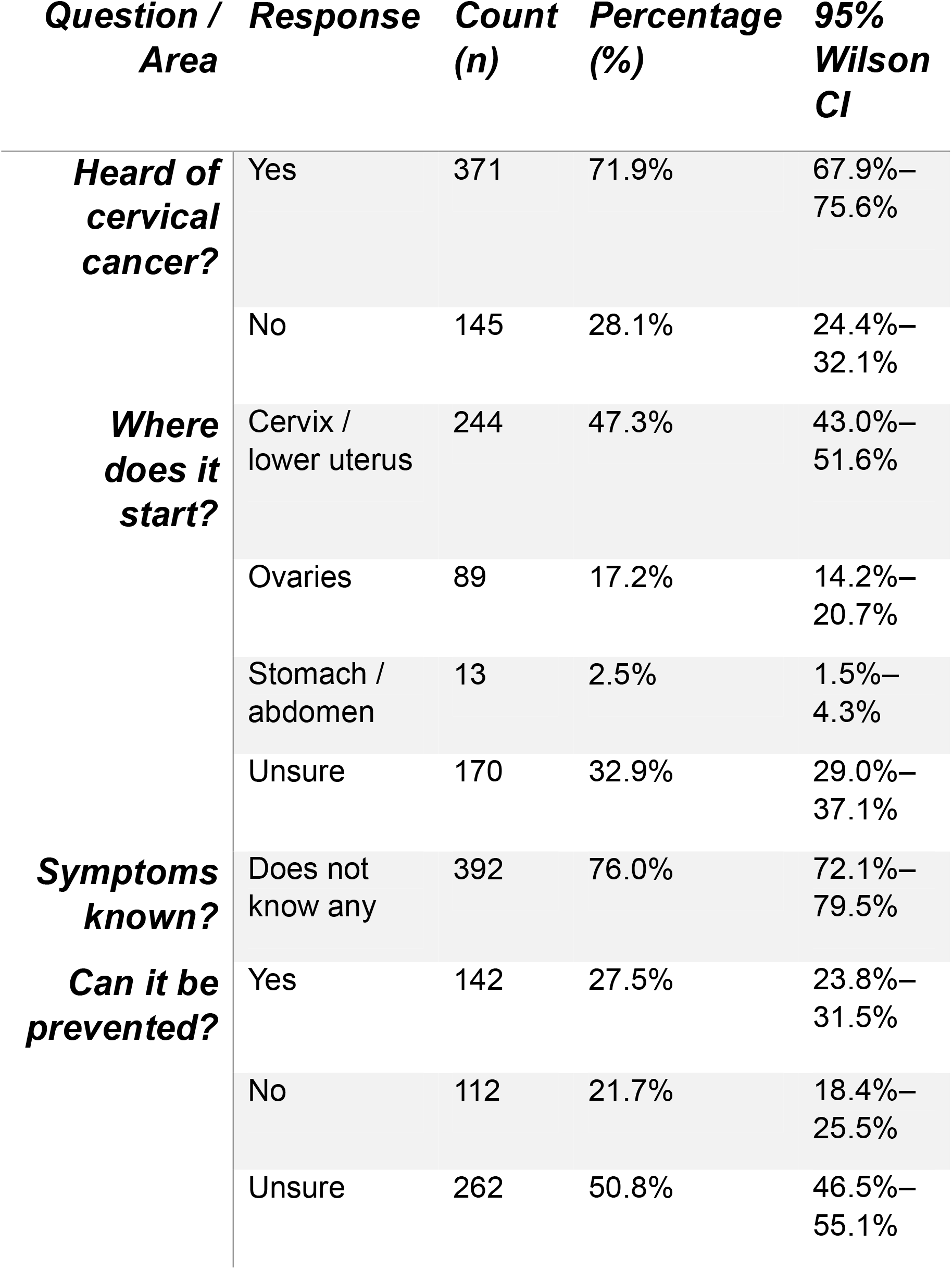
Cervical Cancer Knowledge and Preventability.

### Knowledge About HPV and Transmission

Only 41.9% had heard of HPV. While 53.9% correctly affirmed that HPV causes cervical cancer, 46.1% answered incorrectly or were not sure. Direct skin-to-skin contact was recognized as the transmission route by 56.0%, while 44.0% were unsure or believed it spreads through coughing or contaminated water.

**Table 3.** Knowledge of HPV and Transmission Routes.

| **Question / Area** | **Response** | **Count (n)** | **Percentage (%)** | **95% Wilson CI** |
| --- | --- | --- | --- | --- |
| **Heard of HPV?** | Yes | 216 | 41.9% | 37.7%–46.2% |
|  | No | 300 | 58.1% | 53.8%–62.3% |
| **Does HPV cause cervical cancer?** | True | 278 | 53.9% | 49.6%–58.1% |
|  | False | 126 | 24.4% | 20.9%–28.3% |
|  | Unsure | 112 | 21.7% | 18.4%–25.5% |
| **How does it spread?** | Skin-to-skin / sexual contact | 289 | 56.0% | 51.7%–60.2% |
|  | Airborne / coughing | 51 | 9.9% | 7.6%–12.8% |
|  | Food or water | 19 | 3.7% | 2.4%–5.7% |
|  | Unsure | 157 | 30.4% | 26.6%–34.5% |

### Vaccine Understanding and Uptake

While 61.6% had heard of the HPV vaccine, only 50.0% knew it prevents infection. Nearly one in five (19.8%) mistakenly believed it treats established cancer. Personal vaccine uptake was 12.8% (66/516), but 64 of those 66 students had not completed their schedule. Only two girls (0.4% of all participants) were fully vaccinated.

**Table 4.**
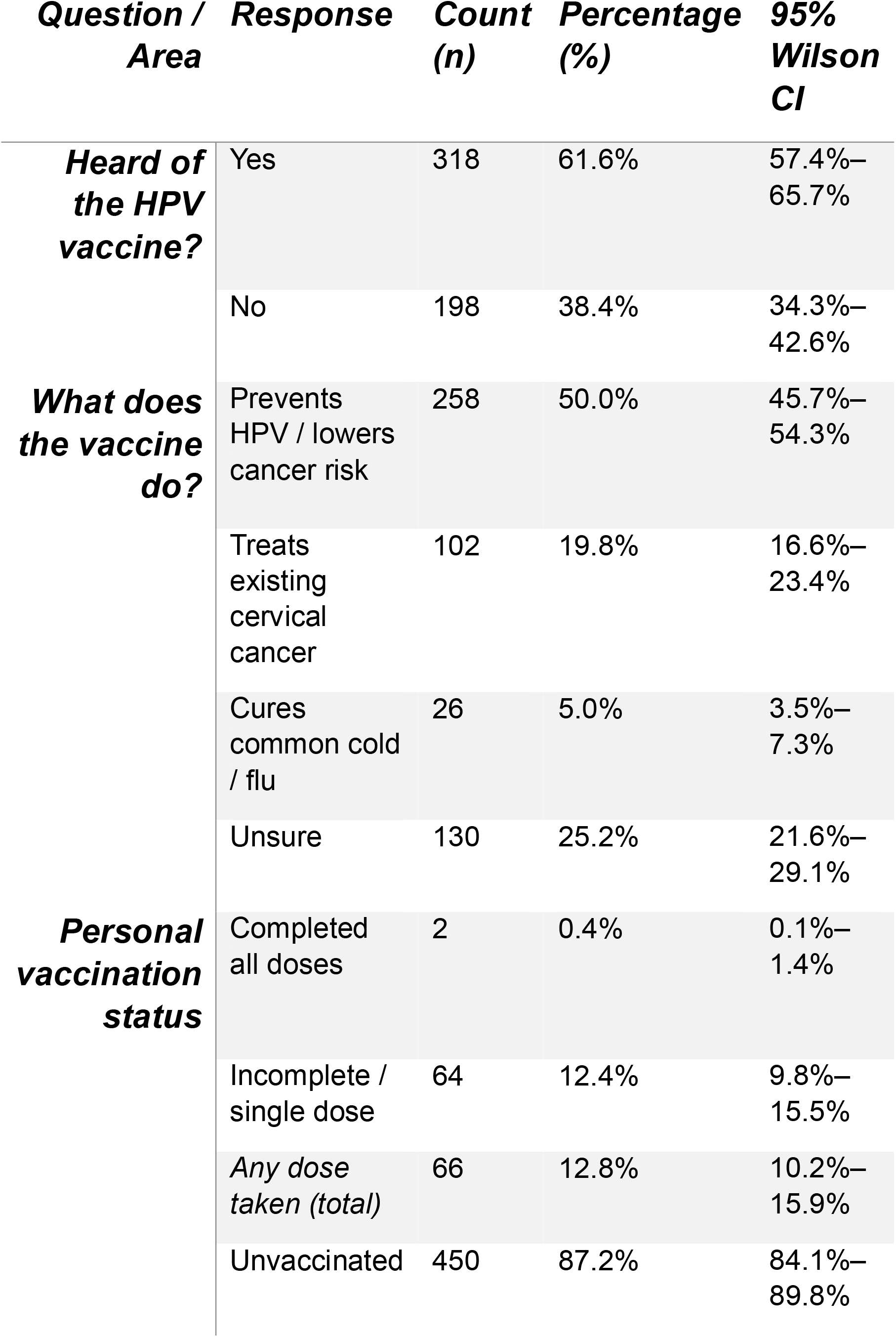
Vaccine Knowledge and Vaccination Status.

### Student Decisions and Rumours

When asked if they would get vaccinated upon a doctor’s recommendation, 45.3% stated they needed more information first. This group was significantly larger than the group that refused (34.1%) or agreed immediately (20.5%). In addition, 75.6% held active worries or uncertainty about whether the vaccine causes infertility, and 96.5% had never heard of a Pap smear.

**Table 5.**
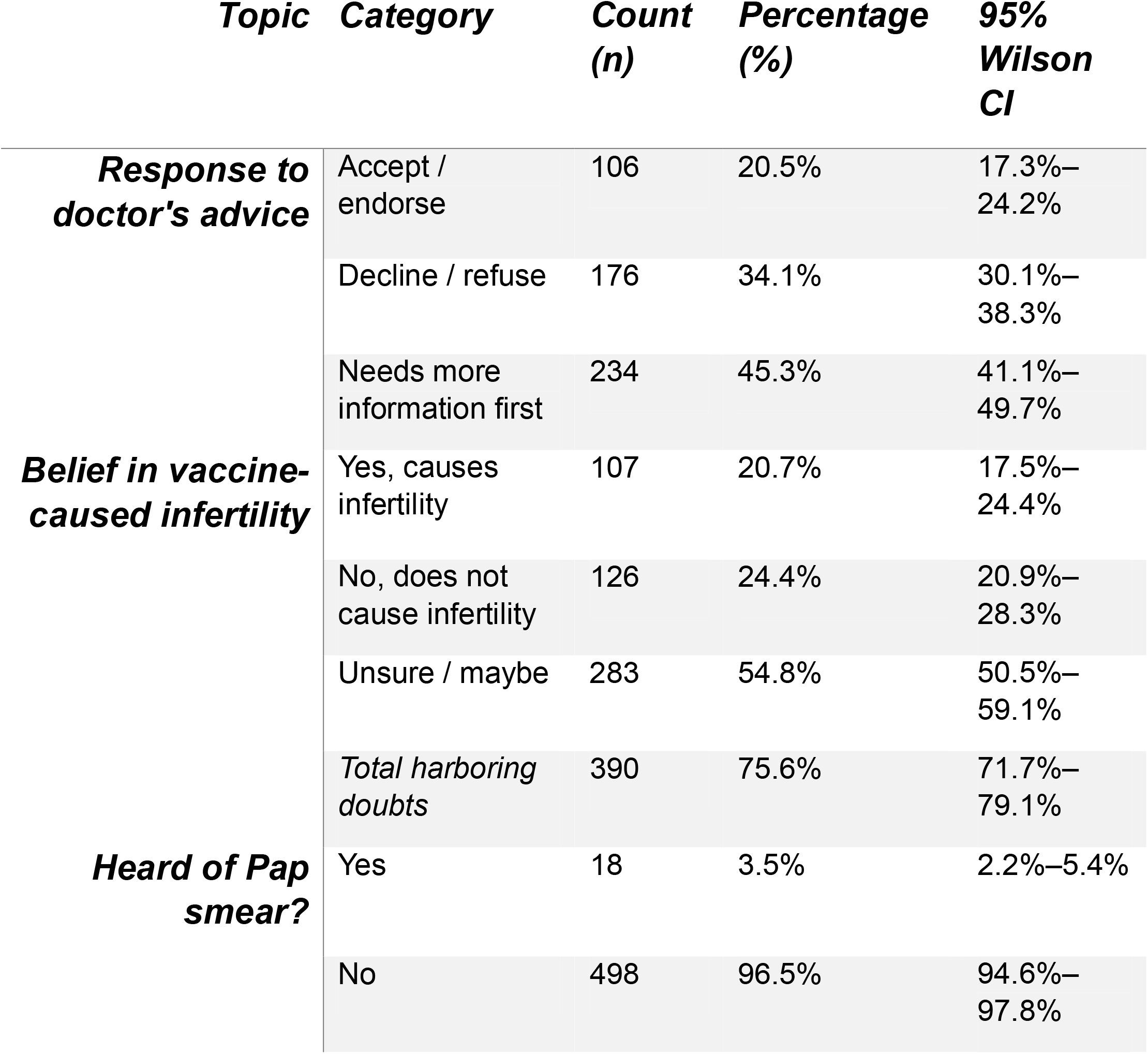
Decisions and Concerns.

## Discussion

These results show a clear gap between hearing the name of a health condition and understanding what to do about it. While most girls had heard of cervical cancer (71.9%) and the vaccine (61.6%), functional knowledge was low. The sharpest drops occurred between hearing about cancer and knowing it is preventable (44.4% drop), hearing about the vaccine and getting vaccinated (48.8% drop), and starting the vaccine series versus finishing it (only 3.0% of recipients finished). In addition, three out of four girls had doubts about infertility, and 45.3% deferred their choice until they received more facts.

This divide matches observations across India [5], [6]. Surface-level awareness comes from brief slogans that fail to explain how diseases develop. When 76.0% of students cannot identify a single symptom and half do not know the disease is preventable, they may view cancer as unavoidable, making them less likely to seek protection. Health campaigns must move beyond slogans and clearly emphasize that cervical cancer can be prevented.

The gap between vaccine awareness (61.6%) and HPV awareness (41.9%) explains why many students misunderstand the vaccine’s purpose. When students and parents do not know that the vaccine must be given before exposure to the virus, they do not see why it should be taken during early adolescence. Clear explanations of transmission and preventive protection are necessary.

Importantly, our data shows that most unvaccinated girls are not opposed to vaccines. The largest single group (45.3%) simply wanted more information before deciding. Focused classroom sessions can double vaccine acceptance [7]. Giving out consent forms on the day of vaccination without prior explanation often leads families to decline out of caution.

Fears about infertility (75.6%) represent a major barrier. In South Asia, rumors that vaccines harm reproduction have disrupted past campaigns for tetanus and polio [10], [11] as well as earlier HPV demonstration projects [12]. Avoiding the topic makes families suspicious. Health teams must directly reassure communities using safety data from hundreds of millions of doses administered worldwide [13], [14]. Finally, teaching older students about Pap smears establishes a healthy foundation for adult screening, supporting the WHO’s dual strategy of vaccination and screening [1], [4].

## Data Availability

All data produced in the present work are contained in the manuscript

